# School masking and COVID-19 community transmission: a synthetic control study

**DOI:** 10.1101/2025.08.11.25333446

**Authors:** Xianqun Luan, Brian T Fisher, Susan E Coffin, David Rubin, Meredith Matone, Jing Huang

**Affiliations:** PolicyLab, Children’s Hospital of Philadelphia; Division of Infectious Disease, Children’s Hospital of Philadelphia; Department of Pediatrics, Perelman School of Medicine, The University of Pennsylvania; University of California, California, USA; Department of Biostatistics, Epidemiology, and Informatics, Perelman School of Medicine, The University of Pennsylvania

## Abstract

**Background:** K-12 schools play a crucial role not only as educational settings but also as hubs for social interaction among children, making them potential drivers of disease transmission within families and the broader community. Little is known about the impact of mandatory school masking policies on community SARS-CoV-2 infection rates during the COVID-19 pandemic. This study addressed this gap by evaluating the association between mandatory school masking policies and community infection rates, accounting for temporal and regional variations.

**Methods:** We conducted a retrospective quasi-experimental study using the synthetic control method. The study focused on the fall 2021 school reopening period in the United States, a time when most U.S. schools returned to fully in-person learning but accompanied by substantial variation in masking policies. Analyses controlled community characteristics prior to reopening and baseline infection rates.

**Findings:** Counties with mandatory school masking policies experienced lower SARS-CoV-2 infection rates compared to counties with non-mandatory policies. During the first nine weeks after school reopening, there was a reduction of 1,096 cases per 100,000 people (95% confidence interval: 880 to 1,310 fewer cases per 100,000 people). This association was influenced by baseline infection rates, population density, and mobility patterns.

**Interpretation:** Mandatory school masking policies were associated with notable reductions in community SARS-CoV-2 infection rates during the fall 2021 school reopening period. This study highlights the potential role of school masking as a public health intervention in mitigating community transmission.

**Funding:** Centers for Disease Control and Prevention and the National Institutes of Health

## Introduction

Kindergarten through 12th grade (K-12) schools are unique environments with extensive human interaction, where children closely interact with peers and then return home to engage with their families. This dynamic creates a potential pathway for infectious diseases, like SARS-CoV-2, to spread not only within schools but also into the surrounding community. Masking policies in schools were implemented as a critical non-pharmaceutical intervention during the COVID-19 pandemic, aiming to reduce transmission in these high-contact settings. While numerous studies have examined the effects of school masking policies on secondary transmission within school environments and the impact of community masking policy on community SARS-CoV-2 incidence rates, the relationship between mandatory masking policies in public schools and broader community transmission has been less thoroughly investigated.^1-4^

In this study, we aim to examine whether mandatory school masking policies, compared to non-mandatory policies, influenced community SARS-CoV-2 incidence rates during the Fall of 2021, a period when most schools had returned to in-person learning, and masking policies varied significantly across different regions of the United States.^5^ We also explore how community characteristics, such as population density, socioeconomic factors, and local SARS-CoV-2 incidence rates, may have shaped the effectiveness of these policies.

## Method

### Study design

This retrospective study focused on U.S. counties with a single public school district to reduce within-county variation in school policies and facilitate clearer attribution of outcomes. We adopted county selection criteria developed in a prior study assessing impact of instructional modality on disease transmission,^6^ prioritizing counties with adequate population sizes to ensure stable estimates of community incidence rates. Among the 229 counties that met these criteria, 166 counties were selected to ensure sufficient variation in school masking policies within states or in nearby states. This heterogeneity in exposure was essential for evaluating the effects of school masking policies while accounting for regional similarities in demographics, policy environments, and COVID-19 transmission dynamics. Specifically, the selected counties were located in states where both mandatory and non-mandatory school masking policies were present or in states bordering such states at the start of the Fall 2021 semester. Within this subset, 89 counties implemented mandatory school masking policies, while 77 counties implemented non-mandatory masking policies. For each county, time zero was defined as the start date of the Fall 2021 academic semester. County-level data for the analyses were collected over a 15-week period, spanning six weeks before and nine weeks after time zero (Fall semester 2021 school opening date).

### Intervention

The intervention was the county-level K-12 public school masking policy, categorized as mandatory versus non-mandatory masking. Hereafter, we refer to schools with mandatory school masking policy as *treated* and schools with non-mandatory masking policies as c*ontrol*. Masking policy data were extracted from official documents published on school and district websites and classified as follows: 1) mandatory masking (*treated)*--school districts that required masking at the start of the Fall 2021 semester, and 2) non-mandatory masking (*control)*--schools districts where masking was optional, recommended, or applied in limited settings (e.g., hallways, grade-specific recommendations) at the start of the Fall 2021semester. To ensure the accuracy of masking policy classification, a blinded independent data collection and validation process was conducted for a random 20% sample of school districts, yielding 92% agreement between initial and validation coding.

### Outcome

The main outcome was the weekly SARS-CoV-2 incidence rate per 100,000 residents at the county level. Daily incidence data were sourced from USAFacts, and population size estimates were obtained from the U.S. Census Bureau.^7, 8^ Both confirmed and probable cases were included following CDC guidelines.

### Covariates

County-level covariates were included to account for potential confounding factors, encompassing demographics, health-related variables, mobility data, and vaccination rates. Key data sources included: demographics derived from the American Community Survey Data^8^, including population density, urban population rate, senior population (>=64 years), young population (<=18 years), minority, college education, high school diploma, poverty rate, and Supplemental Security Income (SSI); health-related factors acquired from the County Health Rankings & Roadmaps^9^, including any chronic condition, obesity, diabetes, smoking, and excessive drinking; social vulnerability obtained from the CDC/Social Vulnerability Index (SVI)^10^; and mobility data obtained from Google COVID-19 Community Mobility Reports^11^, with weekly mobility calculated as a 7-day average of daily mobility metrics across various categories, such as visits to retail and recreation, grocery and pharmacy, workplaces, and residential areas and SARS-CoV-2 vaccination rates data obtained from CDC^12^. These covariates were selected a priori based on existing evidence of factors influencing county-level SARS-CoV-2 incidence rates.^13-15^

Given a large number of covariates (25 variables), we conducted a principal component analysis (PCA) of these covariates to achieve a reduced-rank presentation.^16^ The top five principal components (PCs), which collectively accounted for over 77% of the total variance of the covariates, were selected.

### Synthetic control group construction

We evaluated the effect of school masking policies on county-level SARS-CoV-2 incidence rate using the synthetic control method (SCM).^17-20^ For each treated county, a synthetic control was constructed as a weighted combination of all eligible control counties from a donor pool specifically built for the treated county. These control county pools, primarily selected from the same state as the treated county, aimed to provide at least five control counties when available. If fewer than five control counties were available within the same state, the donor pool was expanded to include control counties from geographically adjacent states to account for regional similarities in population characteristics, policy environments, and SARS-CoV-2 transmission dynamics. The resulting donor pool sizes ranged from 7 to 32 counties per treated county. County weights for the synthetic controls were calculated using pre-intervention SARS-CoV-2 outcomes and the top five PCs of county-level covariates to minimize differences between treated counties and their corresponding synthetic controls. The quality of the constructed synthetic controls was assessed by the absolute standardized mean difference (ASMD), with an ASMD threshold of 0.5 or less considered indicative of a well-matched synthetic control.^21^ Additional details on the construction of the synthetic controls are provided in the Supplementary Materials. The resulting synthetic controls made up the synthetic control group.

## Statistical analysis

The impact of the intervention was estimated as the differences in weekly SARS-CoV-2 incidence rate between treated counties and their synthetic counterparts during the nine-week post-intervention period. Cumulative incidence rate over this nine-week period was also calculated. The 95% conference intervals (CIs) were derived using the bootstrap method with 1,000 resamples.

To assess variation in intervention effects, we conducted stratified analyses across several subgroups. Treated counties were categorized into high and low groups for each of the following pre-intervention metrics: cumulative SARS-CoV-2 incidence rates, population density, and community mobility. Median values of each metric within the treated counties during the pre-intervention period were used as cutoffs to define high and low categories. Intervention effects were then summarized for each category and compared across the strata. Analyses were conducted in R (version 4.4.0), using the Synth package.

## Results

The final analysis included 35 treated counties along with their corresponding synthetic controls, constructed from 69 control counties. These treated counties and their synthetic counterparts achieved an ASMD of ≤ 0.5 for pre-intervention covariates and outcomes. A flowchart outlining county selection and synthetic control construction is provided in Supplementary Materials Figure S1. Variation in school masking policies was most pronounced in the southern regions of the US, leading to an overrepresentation of treated and control counties from the South Atlantic and East South-Central regions in the final study sample (Figure 1). Figure S2 shows the geographic locations and corresponding weights of control counties used to construct the synthetic control for each treated county, while Table S1 summarizes the total weights assigned to each control county, indicating their relative contribution to the synthetic control group.

**Figure 1.**
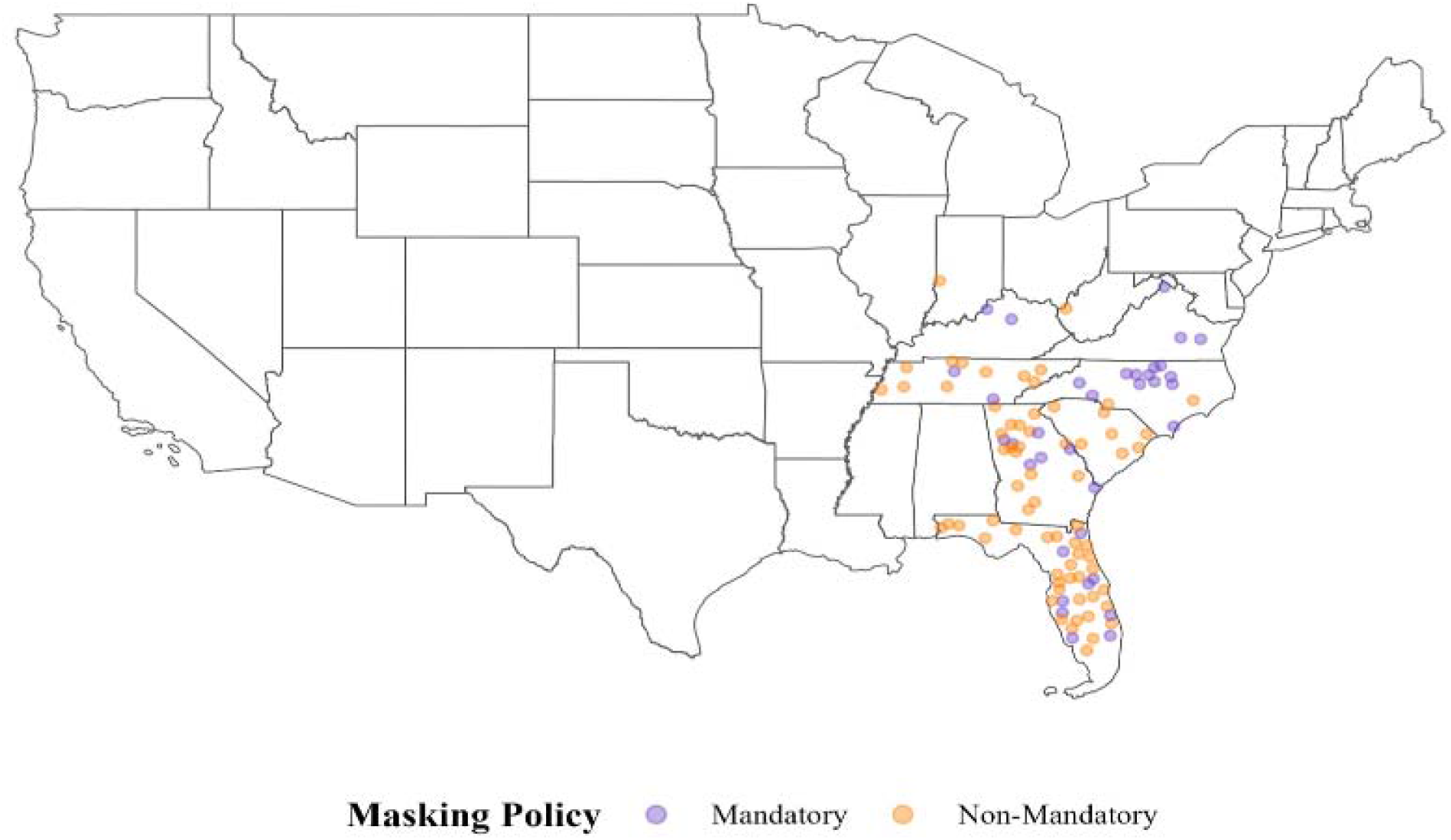
Geographic locations of counties included in the final analysis and their school masking policies at the start of the Fall 2021 semester. This includes 35 treated counties and the 69 control counties used to construct their synthetic controls.

Table 1 shows summary statistics of baseline demographics and SARS-CoV-2 related measures during the pre-intervention period for the 35 treated counties, the 69 control counties used to construct the synthetic controls, and the 35 synthetic controls. Compared to control counties, treated counties had younger populations, were more urban, and had higher population density, higher educational attainment, and higher SARS-CoV-2 vaccination coverage. They also had lower proportions of adults aged 65 or older, lower prevalence of chronic conditions and smoking, lower public insurance coverage, and reduced mobility. The percentage of the population infected with SARS-CoV-2 prior to the start of the Fall 2021 semester was slightly lower in treated counties (10.9% vs. 11.1%). Weekly SARS-CoV-2 incidence rates were also lower in treated counties during the six weeks preceding the semester’s start, with rates of 337.0 vs. 418.0 per 100,000 population in the week immediately before schools opened. These baseline differences were substantially reduced after constructing the synthetic controls. Comparisons of pre-intervention outcomes and county-level covariates between treated counties and their synthetic controls indicate a close balance between the groups, as reflected by small mean differences.

**Table 1.** Summary of pre-intervention outcomes and county-level covariates, reported as mean with standard deviation in parentheses, during the six weeks before the Fall 2021 semester, comparing counties with mandatory vs. non-mandatory school masking policies and mandatory-masking counties vs. their synthetic controls. The 35 treated counties had mandatory masking policies, the 69 control counties had non-mandatory policies, and the 35 synthetic controls were constructed from the control counties.

|  | Treated counties | Control counties | Synthetic controls | Treated vs control counties<sup>#</sup> | Treated vs synthetic controls<sup>#</sup> |
| --- | --- | --- | --- | --- | --- |
| **# Counties** | 35 | 69 | 35 |  |  |
| **Weekly SARS-CoV-2 case rate per 100K before the Fall 2021 semester** |  |  |  |  |  |
| 6th week before school opening (week -5) | 38.3 (29.2) | 41.5 (35.4) | 37.9 (27.6) | -3.2 | 0.4 |
| 5th week before school opening (week -4) | 57.1 (43.0) | 87.2 (147.7) | 56.2 (43.8) | -30.1 | 0.9 |
| 4th week before school opening (week -3) | 200.0 (249.1) | 264.3 (278.7) | 192.9 (248.7) | -64.3 | 7.1 |
| 3rd week before school opening (week -2) | 228.6 (215.8) | 289.2 (252.4) | 228.7 (213.7) | -60.6 | -0.1 |
| 2nd week before school opening (week -1) | 276.6 (180.5) | 333.2 (234.2) | 279.4 (180.6) | -56.6 | -2.8 |
| 1st week before school opening (week 0) | 337.0 (180.1) | 418.0 (258.0) | 340.4 (175.1) | -81 | -3.4 |
| **Population percentage of reported SARS-CoV-2 infection (%)** | 10.9 (1.9) | 11.1 (2.6) | 11.5 (1.8) | -0.2 | -0.6 |
| **Population percentage of COVID-19 vaccination\*** | 38.6 (14.0) | 35.5 (13.0) | 37.0 (10.3) | 3.1 | 1.6 |
| **Population demographics** |  |  |  |  |  |
| Population Density in log scale | 6.3 (0.8) | 5.5 (0.8) | 6.0 (0.7) | 0.8 | 0.3 |
| % Urban population | 82.4 (18.5) | 71.0 (18.8) | 73.2 (11.8) | 11.4 | 9.2 |
| % Population >=64 Years | 16.4 (4.7) | 20.0 (8.1) | 17.1 (3.4) | -3.6 | -0.7 |
| % Population <=18 Years | 21.7 (2.5) | 21.2 (3.7) | 22.6 (2.2) | 0.5 | -0.9 |
| % Minority | 43.0 (16.2) | 29.7 (13.4) | 33.1 (10.9) | 13.3 | 9.9 |
| % College | 32.3 (10.3) | 25.4 (8.6) | 26.6 (4.3) | 6.9 | 5.7 |
| % High school diploma | 11.1 (3.4) | 11.7 (4.7) | 11.9 (2.3) | -0.6 | -0.8 |
| % Poverty | 14.5 (5.3) | 13.8 (5.1) | 13.9 (2.8) | 0.7 | 0.6 |
| Crowding index | 2.1 (0.7) | 1.9 (1.0) | 2.1 (0.5) | 0.2 | 0 |
| CDC Social Vulnerability Index (SVI) | 58.9 (25.5) | 54.8 (26.9) | 55.0 (15.5) | 4.1 | 3.9 |
| Supplemental Security Income (SSI) | 27.4 (9.7) | 27.4 (9.0) | 26.0 (4.6) | 0 | 1.4 |
| **Population Chronic and Health** |  |  |  |  |  |
| % Any chronic condition | 43.9 (4.9) | 46.2 (4.9) | 45.9 (2.8) | -2.3 | -2 |
| % Obesity | 34.0 (3.5) | 33.7 (4.1) | 34.6 (2.6) | 0.3 | -0.6 |
| % Diabetes | 11.5 (1.9) | 11.2 (1.7) | 11.5 (1.0) | 0.3 | 0 |
| % Smoking | 18.2 (3.0) | 20.0 (2.9) | 19.7 (1.4) | -1.8 | -1.5 |
| % Excessive drinking | 18.2 (2.2) | 19.4 (2.8) | 18.6 (2.2) | -1.2 | -0.4 |
| **Health insurance** |  |  |  |  |  |
| % No health insurance | 10.8 (2.8) | 11.4 (3.3) | 11.1 (2.1) | -0.6 | -0.3 |
| % Public insurance | 34.3 (6.4) | 39.0 (9.3) | 36.1 (4.0) | -4.7 | -1.8 |
| % Private insurance | 67.6 (7.4) | 64.9 (8.6) | 66.2 (4.2) | 2.7 | 1.4 |
| Google community mobility^ (mean percentage change from baseline) |  |  |  |  |  |
| Workplaces | -25.3 (5.5) | -21.9 (6.1) | -22.4 (3.5) | -3.4 | -2.9 |
| Grocery and pharmacy | 5.6 (10.0) | 9.8 (13.9) | 10.0 (9.6) | -4.2 | -4.4 |
| Residential | 5.0 (1.6) | 4.3 (1.4) | 4.0 (1.1) | 0.7 | 1 |
| Retail and recreation | -3.4 (9.5) | 3.0 (17.1) | 3.7 (10.5) | -6.4 | -7.1 |

\*Completed as at least one dose of the COVID-19 mRNA vaccines.
^Average percentage change in visits or time spent at specific location categories compared to a baseline period (January 3 - February 6, 2020), derived from anonymized location data.
#The comparisons are reported as mean differences.

Figure 2a illustrates the trends in average weekly SARS-CoV-2 incidence rates throughout the study period for counties with mandatory school masking policies and their synthetic controls. The nearly identical trends during the pre-intervention period indicate a close alignment between the treated counties and their synthetic controls, supporting the validity of the synthetic control construction. The two trends began to diverge approximately one week after the start of the Fall 2021 semester, with treated counties exhibiting lower weekly incidence rates. The largest differences were observed between the third- and sixth-weeks post-intervention, followed by a gradual convergence of the trends by week nine. Figure 2b shows the estimated impact of the mandatory school masking policy on county-level weekly SARS-CoV-2 incidence rates. The effect ranged from 21 fewer weekly cases per 100,000 population in week one (95% CI: -45.25 to 1.67) to a peak reduction of 156 cases per 100,000 (95% CI: -205.0 to -105.5) in week five after the Fall 2021 semester began. After reaching its peak, the effect size gradually declined between weeks five and nine. By the end of the observation period, the cumulative reduction over the nine-week post-intervention period amounted to 820 fewer cases per 100,000 population (95% CI: -1184.8 to -444.0).

**Figure 2.**
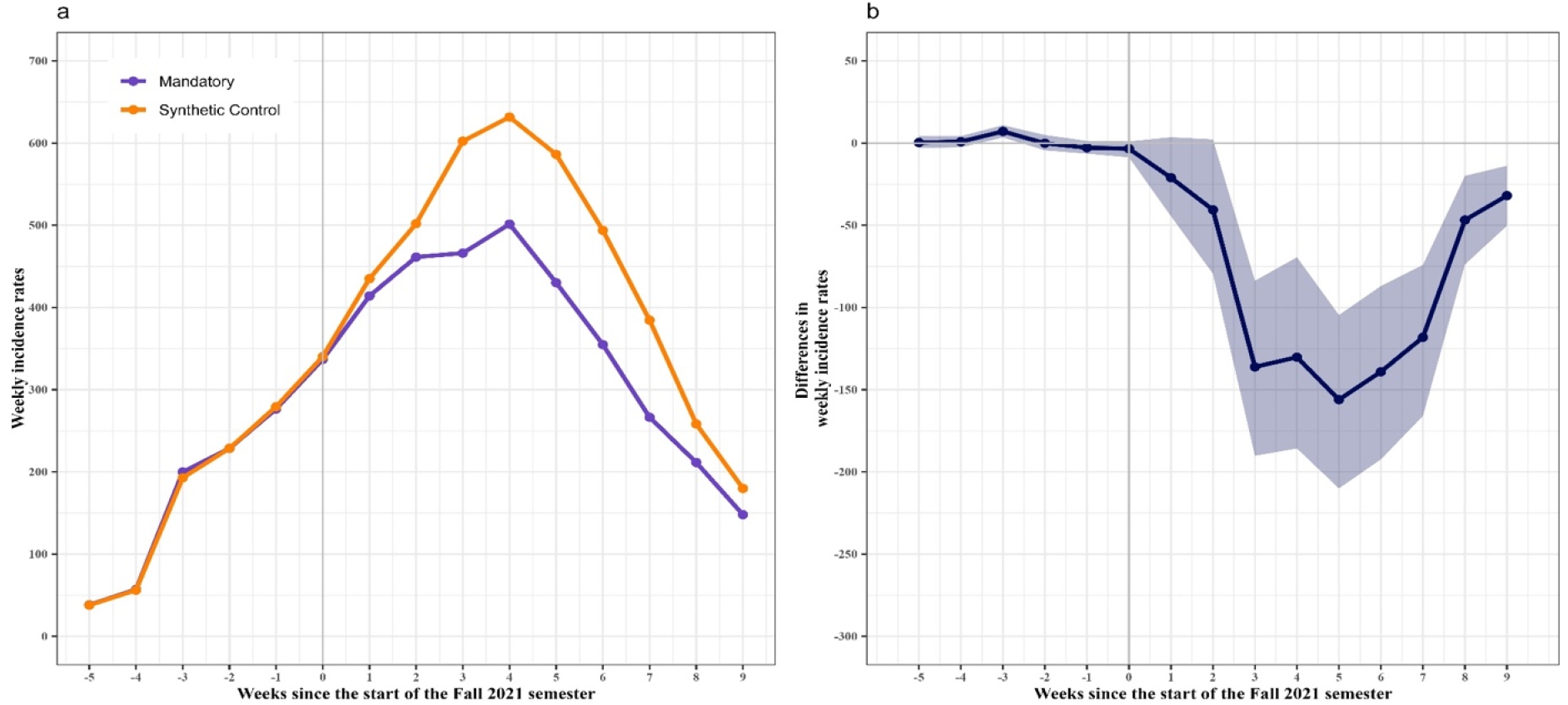
a. Average weekly SARS-CoV-2 incidence rates per 100,000 population in counties with mandatory school masking policies and their synthetic controls during the six weeks before and nine weeks after the start of the Fall 2021 semester. b. Estimated treatment effects of the mandatory school masking policy on county-level SARS-CoV-2 incidence rates per 100,000. Note: The x-axes represent time in weeks during the study period, with zero marking the week that includes the start date of the Fall 2021 semester. Negative values indicate weeks prior to the start date (pre-intervention period), while positive values correspond to weeks following the start date (post-intervention period). The y-axes display the outcomes: in Panel A, the county-level SARS-CoV-2 incidence rate, and in Panel B, the differences in incidence rates between counties with mandatory school masking policies and their synthetic controls. The lines show the mean values, and the shaded areas represent the 95% confidence intervals.

Similar trends in outcomes and intervention effects were observed across subgroups, with stronger effects observed in counties with lower prior population SARS-CoV-2 infection percentages, lower population density, and higher community mobility during the six weeks before the start of the Fall 2021 semester. The most pronounced effects were observed in counties with prior population SARS-CoV-2 infection percentages below 10%, where the largest weekly reduction occurred at week four, with an estimated decrease of approximately 200 fewer weekly cases per 100,000 population (95% CI: -310.8 to -100.5). The cumulative reduction over the nine-week post-intervention period reached 1,031 fewer cases per 100,000 population (95% CI: -1668.5 to -353.6) (Figures 3a&3b). In contrast, counties with higher population SARS-CoV-2 infection percentages before the intervention experienced a less pronounced effect, with a cumulative reduction of 695 cases per 100,000 population over the same period (Figures 3c&3d). Results from analyses stratified by population density and community mobility are presented in Supplemental Figures S3 and S4.

**Figure 3.**
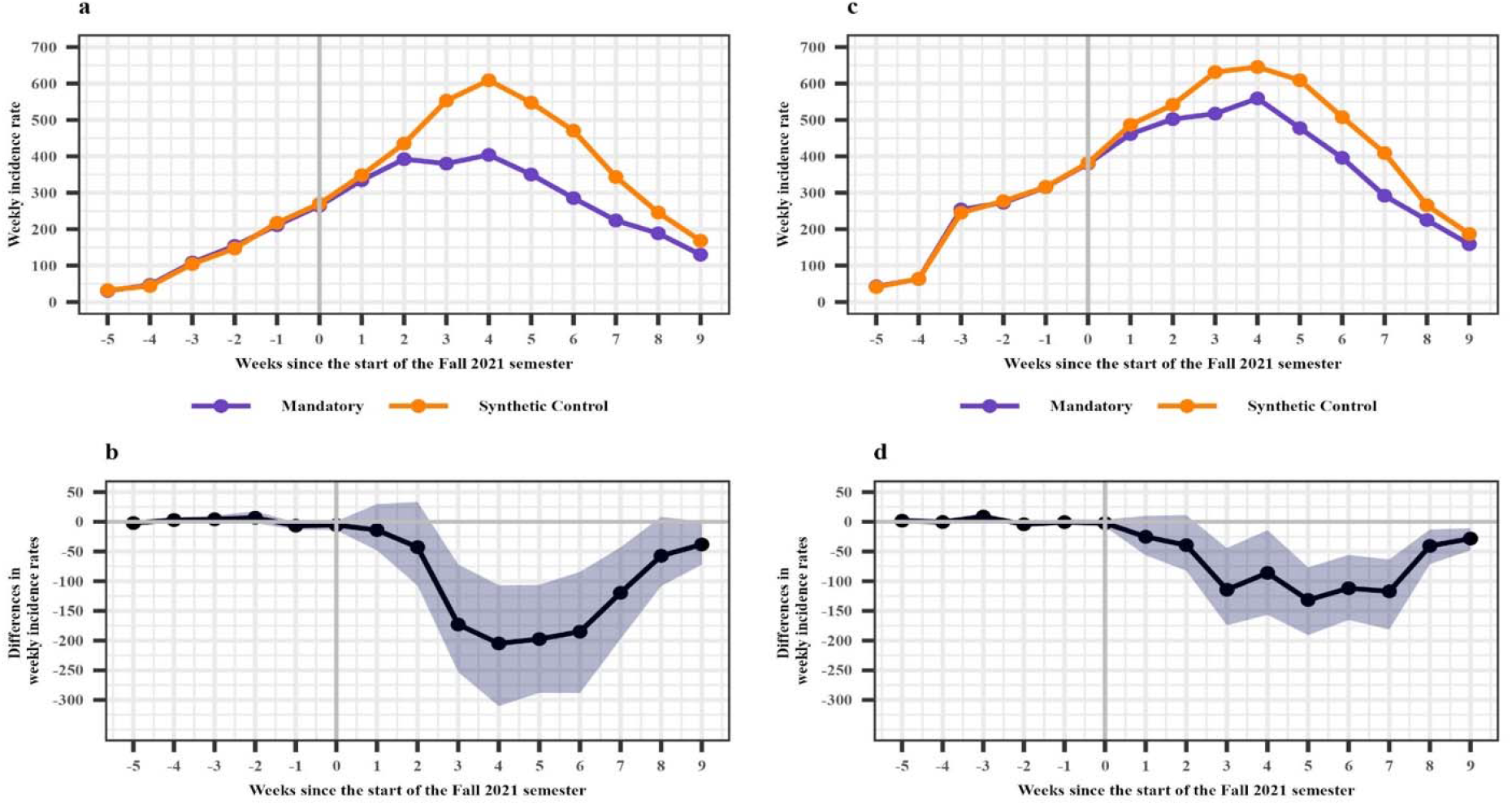
Results of stratified analysis by pre-semester SARS-CoV-2 infection levels: weekly incidence trends and estimated effects of mandatory school masking. Panels a and b: Counties with <10% prior infection; Panels c and d: ≥10%. Panels a and c show average weekly incidence trends per 100,000 between treated and their synthetic controls; panels b and d show treatment effects on incidence per 100,000.

## Discussion

This study employs a rigorous synthetic control study design to compare mandatory versus non-mandatory school masking policies, providing robust evidence that mandatory school masking during the Fall 2021 semester were associated with a significant reduction in community SARS-CoV-2 incidence rates. Counties with mandatory school masking policies experienced lower weekly incidence rates compared to their synthetic controls, with the effect peaking around five weeks into the semester and diminishing by week nine. By the end of the nine-week post-intervention period, the cumulative reduction in SARS-CoV-2 incidence reached 820 fewer cases per 100,000 population. These findings underscore the potential of school masking policies to mitigate the spread of SARS-CoV-2 at the community level.

Our results align with prior studies that have highlighted the role of non-pharmaceutical interventions (NPIs) in controlling community transmission of SARS-CoV-2. However, while many studies have focused on the effects of community-level masking or the reduction of secondary transmission within schools, fewer have investigated the broader impact of school-specific masking policies on community transmission. This study fills that gap by providing evidence of a direct relationship between masking policies in K-12 schools and broader community outcomes, particularly during a time when in-person learning resumed across the United States.

Our subgroup analyses revealed that the effectiveness of school masking policies varied based on community characteristics. The intervention effect was most pronounced in counties with lower pre-intervention SARS-CoV-2 infection percentages, where the cumulative reduction over nine weeks reached 1,031 cases per 100,000 population. This suggests that counties with larger immunological naïve population may benefit more from school masking policies, possibly due to reduced baseline community spread and greater capacity to mitigate surges. Additionally, the intervention effect was greater in counties with lower population density where K-12 schools may represent a larger proportion of total community interactions and in counties with higher community mobility where reducing school transmission may have a broader impact due to frequent movement in the community. These findings highlight the importance of contextual factors in shaping the effectiveness of NPIs and align with prior research suggesting that demographic, social, and behavioral characteristics can modify the impact of public health interventions.

The observed reduction in community incidence rates associated with mandatory school masking policies supports the inclusion of such interventions in broader strategies to control infectious disease transmission. However, that finding alone does not impart judgement on whether required masking policies were a necessity at the time. Policymakers should consider the context-specific effectiveness of school masking policies, particularly in areas with lower pre-existing transmission or other favorable community characteristics. Furthermore, the diminishing effect observed after week five may likely reflect the natural decline in SARS-CoV-2 transmission during the fall wave, rather than a waning impact of masking policies. This suggests that while school-based masking may contribute to short-term reductions in transmission, its effects may align with broader epidemic trends. To sustain low transmission over time, complementary strategies such as vaccination campaigns and expanded testing remain important components of a comprehensive public health response.

A key strength of this study is the use of the SCM, which allowed us to account for unmeasured confounding by creating well-matched synthetic controls for treated counties. The high degree of pre-intervention alignment between treated and synthetic control counties supports the validity of our results. Additionally, the inclusion of a broad range of county-level covariates and stratified subgroup analyses provides a nuanced understanding of how community characteristics influence the effectiveness of school masking policies. However, this study has several limitations. First, the overrepresentation of counties from southern U.S. regions in our final sample may limit the transportability of our findings to other regions with different sociodemographic characteristics or policy environments. Second, the reliance on publicly available data may introduce misclassification bias in masking policies, although our validation process showed high consistency in exposure classification. Third, while the synthetic control method minimizes confounding, unmeasured factors specific to treated counties may still influence the results.

Future studies should explore the long-term effects, including possible negative impact on learning, of school masking policies and their interaction with other NPIs, such as vaccination and testing strategies. Additionally, investigations into the differential impact of masking policies on specific subpopulations, such as students, teachers, and families, could provide insights into more targeted policy development.

## Contributors

XL, DR, MM, and JH conceptualized the study. XL conducted the data analysis with oversight from JH. BTF, SEC, DR, and MM provided input on study design and interpretation of results. XL and JH drafted the manuscript. All authors critically reviewed and revised the manuscript and approved the final version.

## Data sharing

Data for this study are available upon reasonable request to the corresponding author.

## Acknowledgments

This project was supported in part by a grant from the U.S. Centers for Disease Control and Prevention (CDC) under award number U01CK000674. The findings and conclusions in this report are those of the authors and do not necessarily represent the official position of the CDC.

## Declaration of generative AI and AI-assisted technologies in the writing process

During the preparation of this work the authors used chatgpt 4o to improve language and readability. After using this tool, the authors reviewed and edited the content as needed and take full responsibility for the content of the publication.

## References

1. Boutzoukas AE, Zimmerman KO, Inkelas M, Brookhart MA, Benjamin Sr DK, Butteris S, et al. School masking policies and secondary SARS-CoV-2 transmission. Pediatrics. 2022;149(6):e2022056687.

2. Cowger TL, Murray EJ, Clarke J, Bassett MT, Ojikutu BO, Sánchez SM, et al. Lifting universal masking in schools—Covid-19 incidence among students and staff. New England Journal of Medicine. 2022;387(21):1935–46.

3. Donovan CV. SARS-CoV-2 incidence in K–12 school districts with mask-required versus mask-optional policies—Arkansas, August–October 2021. MMWR Morbidity and mortality weekly report. 2022;71.

4. Moorthy GS, Mann TK, Boutzoukas AE, Blakemore A, Brookhart MA, Edwards L, et al. Masking adherence in K–12 schools and SARS-CoV-2 secondary transmission. Pediatrics. 2022;149(Supplement_2).

5. Camera L. Nearly 100% of Students Back to School Full Time and In Person. US News. 2021.

6. Matone M, Wang X, Marshall D, Huang J, Worsley D, Filograna C, et al. Association of in-person vs virtual education with community COVID-19 case incidence following school reopenings in the first year of the COVID-19 pandemic. JAMA Network Open. 2023;6(4):e238300–e.

7. USAFacts. US COVID-19 cases and deaths by state 2023 [Available from: https://usafacts.org/visualizations/coronavirus-covid-19-spread-map/.

8. Bureau USC. American community survey data 2020 [Available from: https://www.census.gov/programs-surveys/acs/data.html.

9. Roadmaps CHR. Health data 2020 [Available from: https://www.countyhealthrankings.org/.

10. ATSDR. Place and health - geospatial research, analysis, and services program 2020 [Available from: https://www.atsdr.cdc.gov/placeandhealth/svi/index.html.

11. Google. COVID-19 community mobility reports 2020 [Available from: https://www.google.com/covid19/mobility/.

12. CDC. COVID-19 vaccinations in the United States, county 2021 [Available from: https://dev.socrata.com/foundry/data.cdc.gov/8xkx-amqh.

13. McLaughlin JM, Khan F, Pugh S, Angulo FJ, Schmitt H-J, Isturiz RE, et al. County-level predictors of coronavirus disease 2019 (COVID-19) cases and deaths in the United States: what happened, and where do we go from here? Clinical Infectious Diseases. 2021;73(7):e1814–e21.

14. Khan SS, Krefman AE, McCabe ME, Petito LC, Yang X, Kershaw KN, et al. Association between county-level risk groups and COVID-19 outcomes in the United States: a socioecological study. BMC Public Health. 2022;22:1–9.

15. Rubin D, Huang J, Fisher BT, Gasparrini A, Tam V, Song L, et al. Association of social distancing, population density, and temperature with the instantaneous reproduction number of SARS-CoV-2 in counties across the United States. JAMA network open. 2020;3(7):e2016099–e.

16. Bishop CM, Nasrabadi NM. Pattern recognition and machine learning: Springer; 2006.

17. Abadie A, Gardeazabal J. The economic costs of conflict: A case study of the Basque Country. American economic review. 2003;93(1):113–32.

18. Abadie A, Diamond A, Hainmueller J. Synthetic control methods for comparative case studies: Estimating the effect of California’s tobacco control program. Journal of the American statistical Association. 2010;105(490):493–505.

19. Bouttell J, Popham F, Lewsey J, Robinson M, Craig P. Use of synthetic control methodology for evaluating public health interventions: a literature review. The Lancet. 2017;390:S26.

20. Britteon P, Fatimah A, Lau Y-S, Anselmi L, Turner AJ, Gillibrand S, et al. The effect of devolution on health: a generalised synthetic control analysis of Greater Manchester, England. The Lancet Public Health. 2022;7(10):e844–e52.

21. Parast L, Hunt P, Griffin BA, Powell D. When is a match sufficient? a score-based balance metric for the synthetic control method. Journal of Causal Inference. 2020;8(1):209–28.

