## Supplementary Materials for "School masking and COVID-19 community transmission: a synthetic control study"

**Additional details on the construction of synthetic controls**

**Donor pool:** The donor pool for constructing synthetic controls was designed to ensure regional comparability and adequate representation for each treated county. Counties in the donor pool, referred to as donor counties, served as the building blocks for creating synthetic controls. Initially, donor counties were selected primarily from the same state as the treated county to account for similarities in population characteristics, policy environments, and SARS-CoV-2 transmission dynamics. This state-based approach preserved regional context, which is critical for maintaining comparability between treated and control counties. To ensure a sufficient number of donor counties, the donor pool was expanded to include counties from geographically adjacent states if fewer than five eligible donor counties were available within the same state. This expansion allowed for the inclusion of additional comparable counties while maintaining regional similarities. The resulting donor pool sizes ranged from 7 to 32 counties per treated county. Weighted combinations of donor counties were used to closely match pre-intervention SARS-CoV-2 outcomes and key covariates for the treated county. This systematic approach ensured robust and well-matched synthetic controls for the analysis.

**Synthetic control construction:** After the donor pool was constructed, synthetic controls were created using the R package Synth. For each treated county, a synthetic control was developed as a weighted combination of donor counties to closely match the pre-intervention characteristics of the treated county. Key matching variables included baseline SARS-CoV-2 weekly incidence rates (six variables each for a week during the pre-intervention period) and the top five principal components (PC1-PC5) of county-level covariates, capturing population characteristics. Loadings of each characteristic on these five principal components are shown in Figure S5. Weights for donor counties were optimized to minimize differences between treated counties and their synthetic controls.

To ensure the quality of the synthetic controls, a placebo analysis was performed for each donor county. In this analysis, synthetic controls were constructed for donor counties as if they were treated. The differences in baseline SARS-CoV-2 weekly incidence rates and auxiliary covariates (PC1–PC5) between the actual treated counties and their synthetic controls were then compared against the distribution of differences observed in the placebo synthetic controls. If the differences for a treated county fell within the 95% confidence interval of the placebo analysis, the synthetic control was deemed well-matched. Additionally, the quality of the synthetic controls was assessed using the absolute standardized mean difference (ASMD), where an ASMD of 0.5 or less was considered indicative of a well-matched control. Treated counties with synthetic controls that did not meet these criteria were excluded from the outcome analysis. This approach ensured the inclusion of only high-quality synthetic controls, enhancing the reliability and validity of the study’s findings.

**Sensitivity Analysis.** Treatment effects for each pair of treated county and its synthetic control is shown in Figure S6**.** To ensure that the estimated treatment effect is not driven by a small subset of counties, we visualized weekly SARS-CoV-2 incidence rates per 100,000 population for each treated county alongside its corresponding synthetic control over the study period. All pairs exhibited strong alignment in pre-intervention trends, further supporting the validity of the synthetic control construction. Post-intervention patterns were consistent across counties and closely resembled the overall average trends, reinforcing the robustness and generalizability of the observed average treatment effect.

We also compare the summary statistics of baseline demographics and SARS-CoV-2 related measures during the pre-intervention period for the 35 treated counties included in the final analysis versus 54 treated counties excluded due to insufficient balance with their synthetic controls (i.e., ASMD > 0.5). The excluded counties were demographically similar to those included but had slightly lower proportions of urban population. The results are shown in Table S2.

**Figure S1.** Study design flowchart.

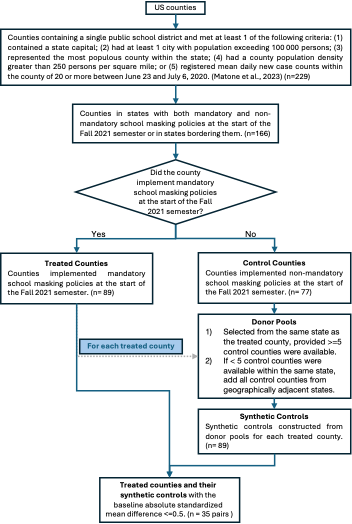

**Figure S2.** Geographic locations and weights of control counties used to construct the synthetic control for each treated county. Treated counties are marked with stars. Control counties are shaded from light yellow to dark purple, with darker colors indicating greater weights in the synthetic control construction.

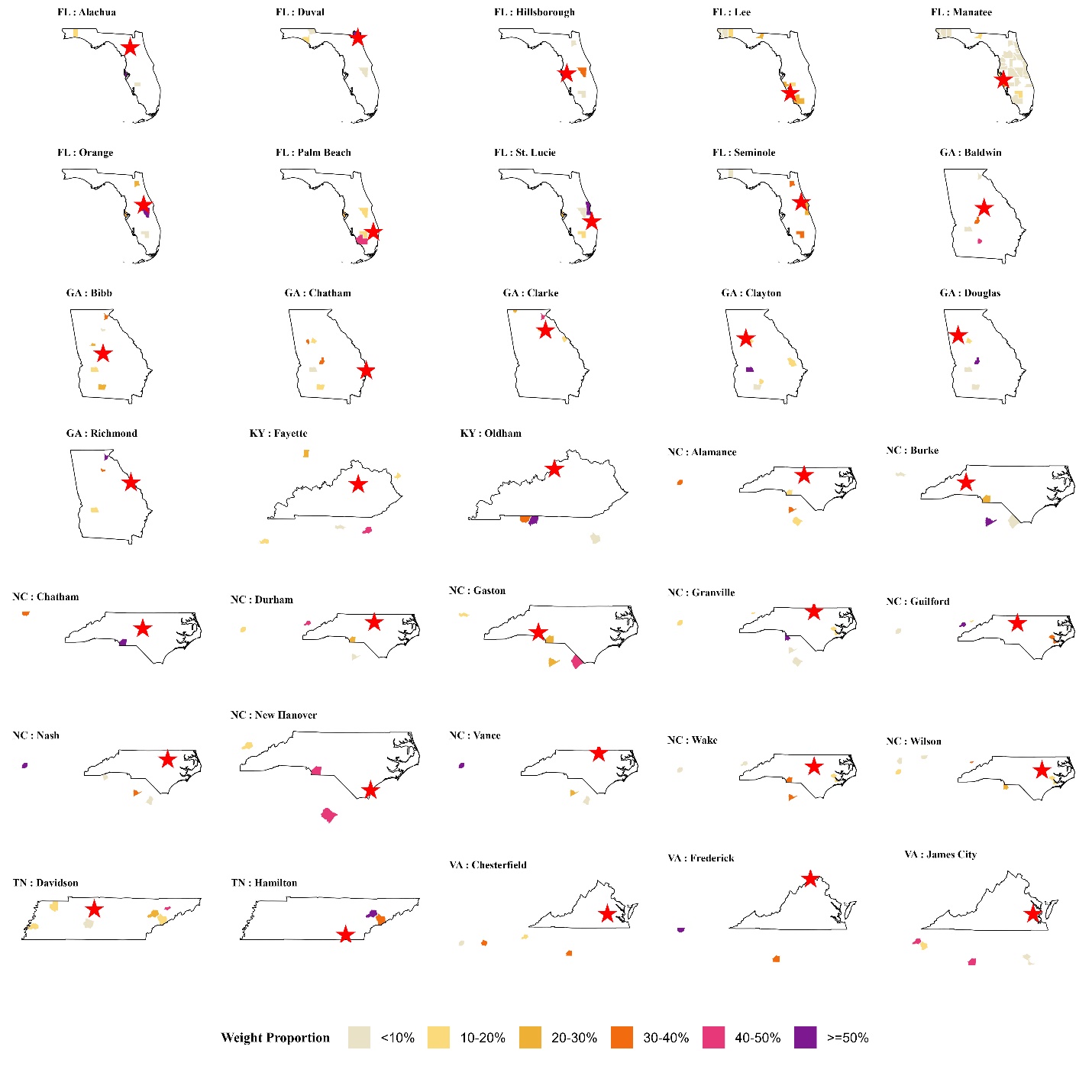

**Table S1.** Total weights assigned to each control county.

| **State** | **County** | **Synthetic Control Total Weights** |
| --- | --- | --- |
| North Carolina | Union County | 3.509 |
| Tennessee | Knox County | 3.135 |
| Tennessee | Madison County | 2.199 |
| Florida | Pinellas County | 2.118 |
| South Carolina | Sumter County | 2.114 |
| Tennessee | Robertson County | 1.441 |
| Georgia | Houston County | 1.390 |
| Georgia | Habersham County | 1.384 |
| Florida | Osceola County | 1.206 |
| Tennessee | Hamblen County | 1.037 |
| Georgia | Sumter County | 1.012 |
| North Carolina | Craven County | 0.869 |
| Florida | Hendry County | 0.801 |
| Florida | Collier County | 0.748 |
| South Carolina | Lancaster County | 0.747 |
| Florida | Brevard County | 0.719 |
| Florida | Nassau County | 0.706 |
| Florida | Clay County | 0.689 |
| Florida | Sarasota County | 0.664 |
| Tennessee | Sevier County | 0.649 |
| Georgia | Tift County | 0.580 |
| Georgia | Barrow County | 0.577 |
| South Carolina | Berkeley County | 0.572 |
| Tennessee | Sumner County | 0.538 |
| South Carolina | Horry County | 0.495 |
| Georgia | Colquitt County | 0.482 |
| Georgia | Henry County | 0.466 |
| Florida | Okaloosa County | 0.370 |
| Tennessee | Maury County | 0.366 |
| Florida | Leon County | 0.357 |
| Georgia | Fayette County | 0.309 |
| Tennessee | Putnam County | 0.306 |
| Georgia | Catoosa County | 0.296 |
| Indiana | Vigo County | 0.260 |
| Georgia | Spalding County | 0.239 |
| Tennessee | Tipton County | 0.231 |
| Florida | DeSoto County | 0.204 |
| Florida | Bay County | 0.182 |
| South Carolina | Georgetown County | 0.177 |
| Georgia | Bulloch County | 0.145 |
| West Virginia | Cabell County | 0.144 |
| Georgia | Columbia County | 0.119 |
| Tennessee | Weakley County | 0.118 |
| South Carolina | Pickens County | 0.091 |
| Florida | Jackson County | 0.040 |
| Florida | Santa Rosa County | 0.021 |
| Florida | Martin County | 0.019 |
| Florida | Escambia County | 0.019 |
| Florida | Indian River County | 0.014 |
| Florida | Polk County | 0.013 |
| Florida | Highlands County | 0.012 |
| Florida | Pasco County | 0.012 |
| Florida | Lake County | 0.010 |
| Florida | Marion County | 0.009 |
| Florida | Citrus County | 0.009 |
| Florida | Hernando County | 0.009 |
| Florida | Flagler County | 0.008 |
| Florida | St. Johns County | 0.008 |
| Florida | Suwannee County | 0.008 |
| Florida | Volusia County | 0.007 |
| Florida | Putnam County | 0.006 |
| Florida | Columbia County | 0.006 |
| Florida | Sumter County | 0.003 |
| South Carolina | Aiken County | 0.003 |
| Florida | Charlotte County | 0.003 |
| Georgia | Forsyth County | 0.001 |
| Georgia | Coweta County | 0.000 |
| Georgia | Cherokee County | 0.000 |
| Georgia | Paulding County | 0.000 |

**Figure S3.** Results of stratified analysis by population density: weekly incidence trends and effects of mandatory school masking. Panels a and b: Population density <400 person/mile^2^; Panels c and d: >=400 person/mile^2^. Panels a and c show average weekly incidence trends per 100,000 between treated and their synthetic controls; panels b and d show treatment effects on incidence per 100,000.

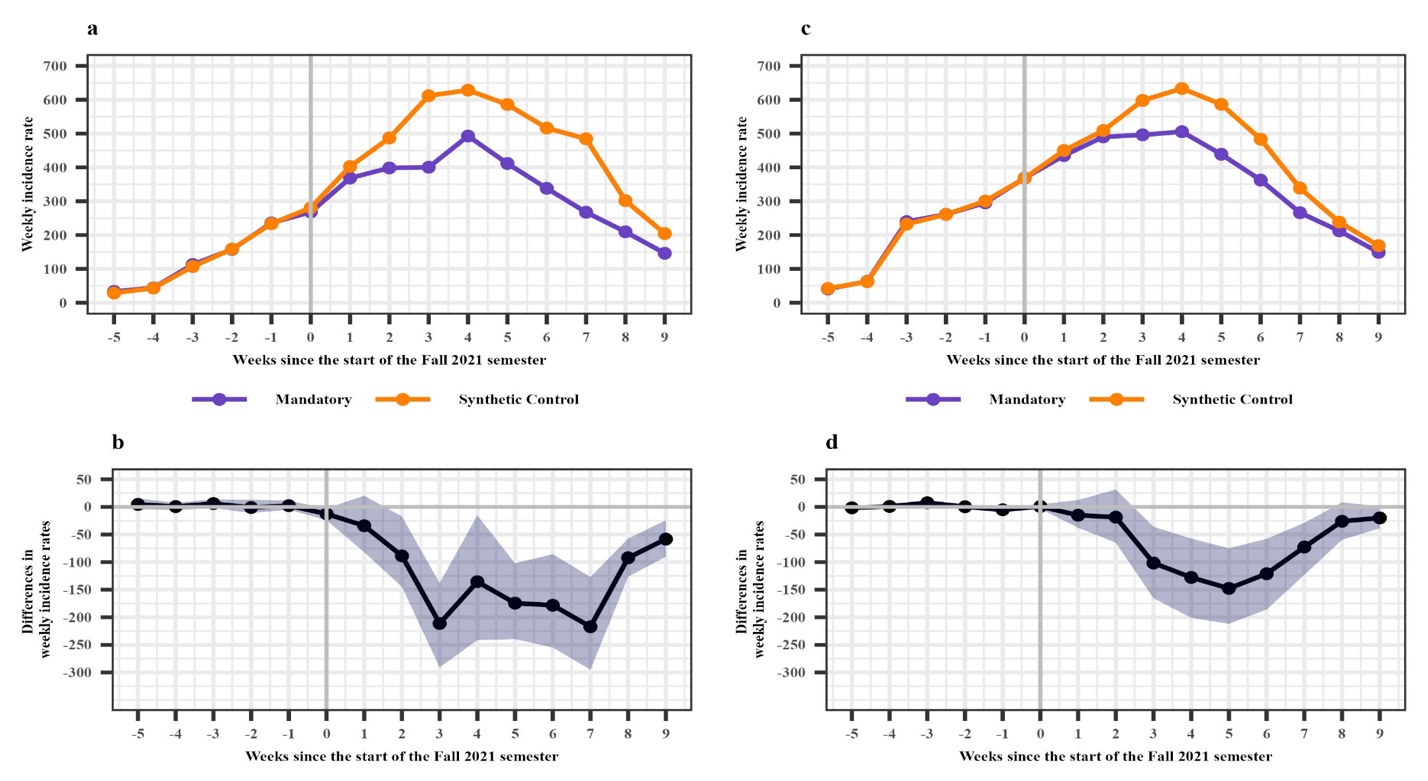

**Figure S4.** Results of stratified analysis by residential mobility change: weekly incidence trends and effects of mandatory school masking. Panels a and b: Mobility increase <5%; Panels c and d: ≥5%. Panels a and c show average weekly incidence trends per 100,000 between treated and their synthetic controls; panels b and d show treatment effects on incidence per 100,000.

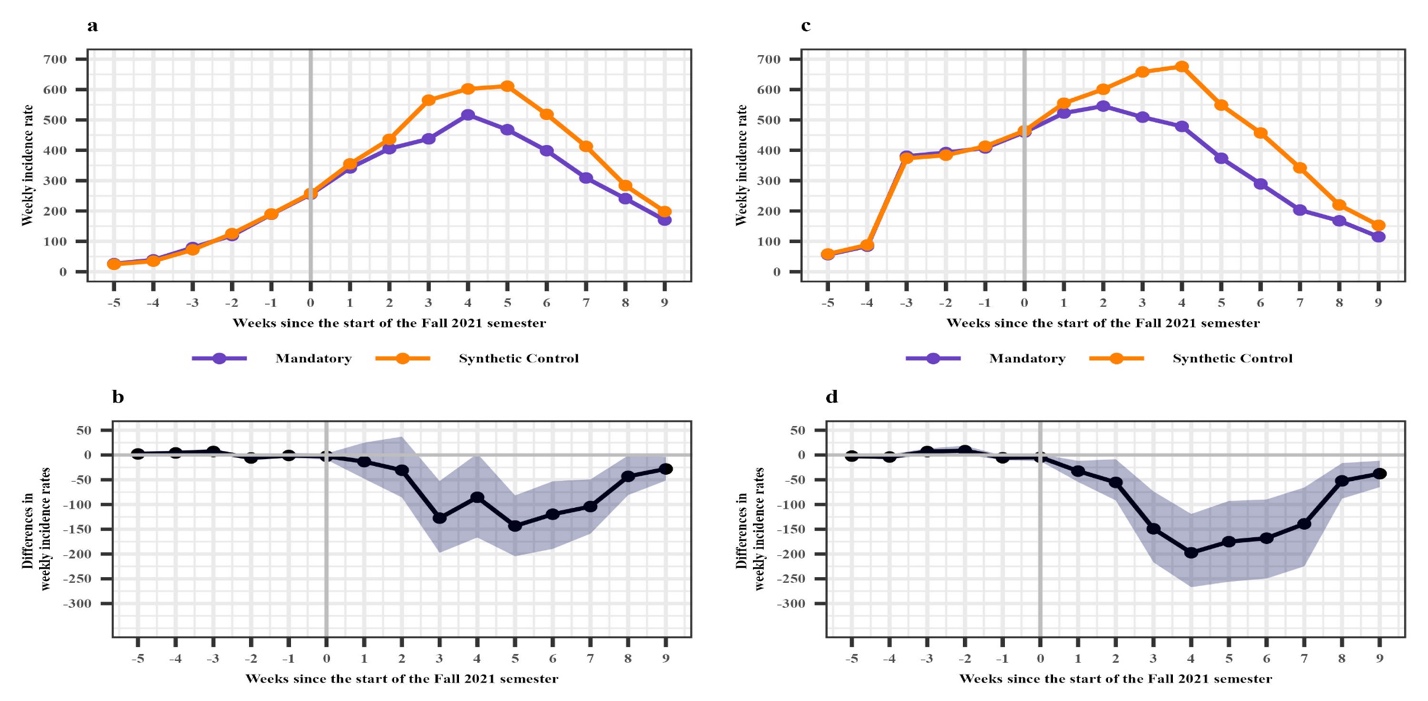

**Figure S5.** Loadings of county-level characteristics on the first five principal components (PCs).
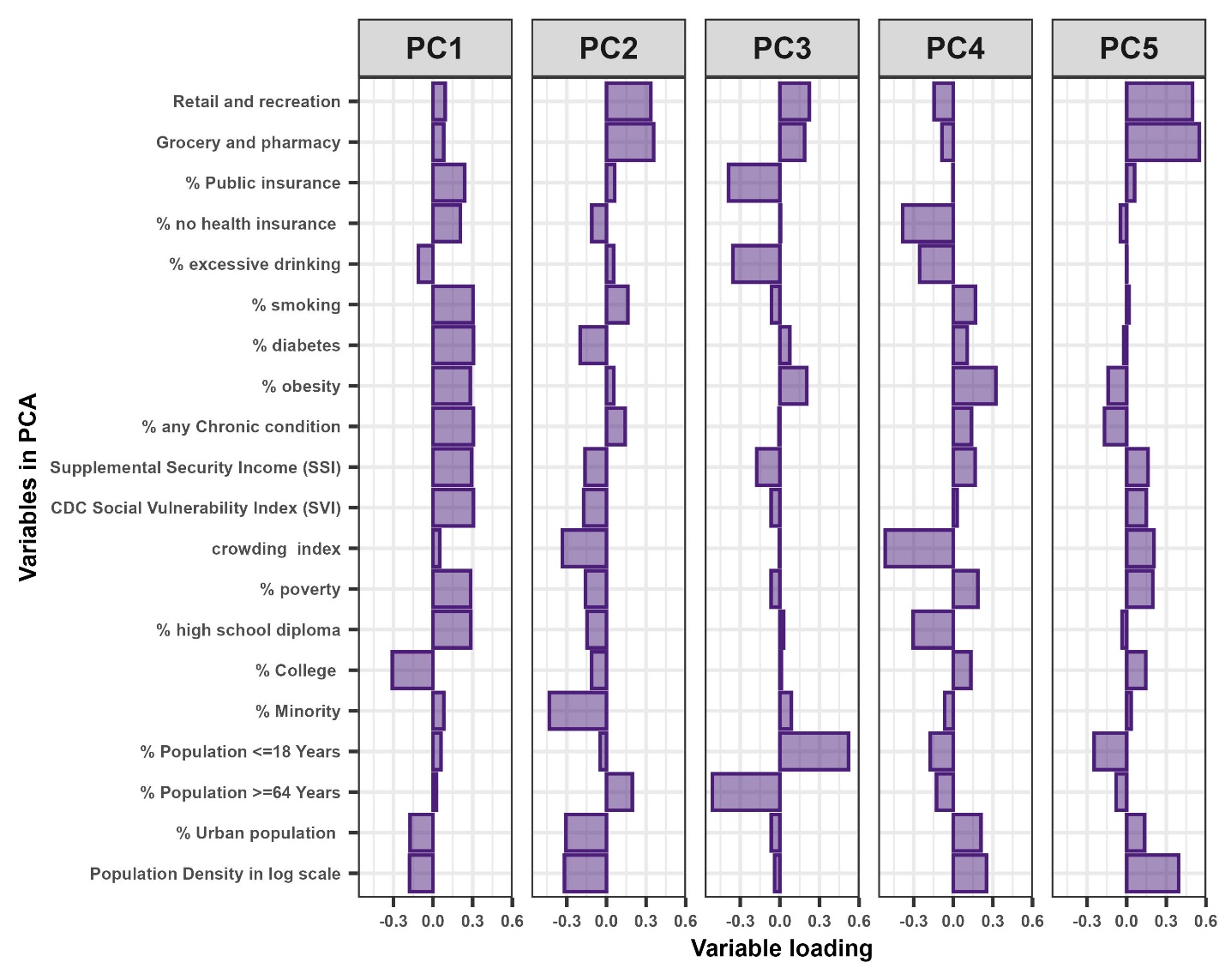

**Figure S6.** Weekly SARS-CoV-2 incidence rates per 100,000 population for each treated county alongside its corresponding synthetic control over the study period.

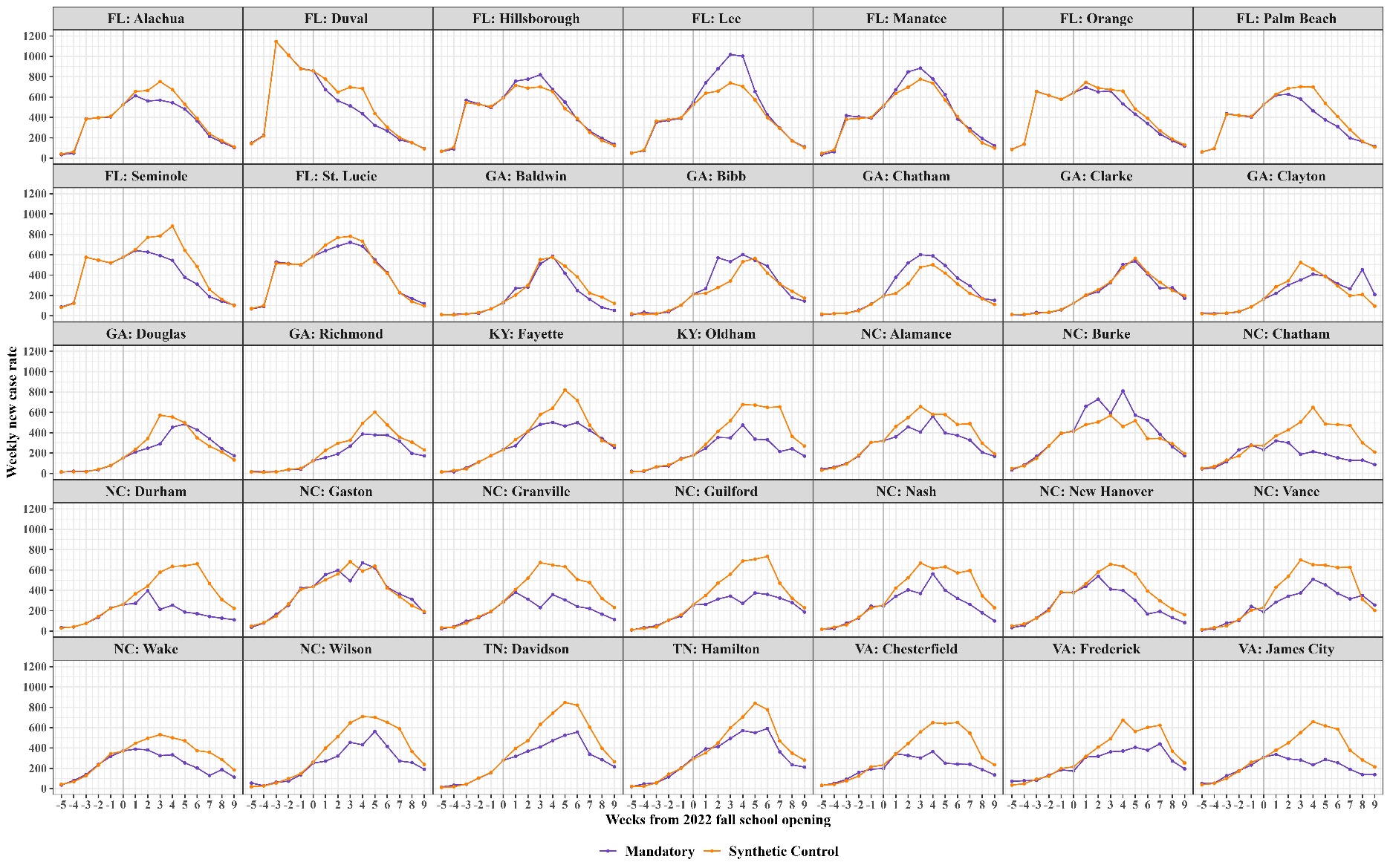

**Table S2.** Summary of pre-intervention outcomes and county-level covariates, reported as mean with standard deviation in parentheses, during the six weeks before the Fall 2021 semester, comparing counties with mandatory school masking policies that were included in the final analysis versus those that were not. Treated counties included are counties with mandatory school masking policies and ASMD of ≤ 0.5 for pre-intervention covariates and outcomes compared to their synthetic controls. Treated counties excluded are counties with mandatory school masking policies and ASMD of > 0.5 for pre-intervention covariates and outcomes compared to their synthetic controls.

|  | **Treated counties included** | **Treated counties excluded** | **Difference^#^** |
| --- | --- | --- | --- |
| **# Counties** | 35 | 54 | 19 |
| **Average weekly SARS-CoV-2 case rate per 100K during the six weeks before the Fall 2021 semester** | 189.6(149.6) | 190.1(151.1) | 0.53 |
| **Population percentage of reported SARS-CoV-2 infection (%)** | 10.9 (1.9) | 10.8 (2.8) | -0.1 |
| **Population percentage of COVID-19 vaccination*** | 38.6 (14.0) | 30.3 (16.6) | -8.3 |
| **Population demographics** |  |  |  |
| Population Density in log scale | 6.3 (0.8) | 6.1 (1.5) | -0.2 |
| % Urban population | 82.4 (18.5) | 69.1 (29.6) | -13.3 |
| % Population >=64 Years | 16.4 (4.7) | 16.0 (4.4) | -0.4 |
| % Population <=18 Years | 21.7 (2.5) | 22.1 (2.8) | 0.4 |
| % Minority | 43.0 (16.2) | 41.2 (19.2) | -1.8 |
| % College | 32.3 (10.3) | 30.5 (14.0) | -1.8 |
| % High school diploma | 11.1 (3.4) | 12.0 (4.3) | 0.9 |
| % Poverty | 14.5 (5.3) | 14.7 (6.2) | 0.2 |
| Crowding index | 2.1 (0.7) | 2.3 (1.2) | 0.2 |
| CDC Social Vulnerability Index (SVI) | 58.9 (25.5) | 57.4 (29.3) | -1.5 |
| Supplemental Security Income (SSI) | 27.4 (9.7) | 27.0 (10.9) | -0.4 |
| **Population Chronic and Health** |  |  |  |
| % Any chronic condition | 43.9 (4.9) | 44.6 (7.4) | 0.7 |
| % Obesity | 34.0 (3.5) | 34.7 (4.8) | 0.7 |
| % Diabetes | 11.5 (1.9) | 11.5 (2.2) | 0 |
| % Smoking | 18.2 (3.0) | 18.7 (4.1) | 0.5 |
| % Excessive drinking | 18.2 (2.2) | 17.7 (2.6) | -0.5 |
| **Health insurance** |  |  |  |
| % No health insurance | 10.8 (2.8) | 10.4 (3.1) | -0.4 |
| % Public insurance | 34.3 (6.4) | 34.9 (8.6) | 0.6 |
| % Private insurance | 67.6 (7.4) | 67.8 (9.5) | 0.2 |
| **Google community mobility^ (mean percentage change from baseline)** |  |  |  |
| Workplaces | -25.3 (5.5) | -23.7 (10.5) | 1.6 |
| Grocery and pharmacy | 5.6 (10.0) | 8.1 (11.3) | 2.5 |
| Residential | 5.0 (1.6) | 4.9 (2.2) | -0.1 |
| Retail and recreation | -3.4 (9.5) | -0.7 (12.8) | 2.7 |

*Completed as at least one dose of the COVID-19 mRNA vaccines.

^ Average percentage change in visits or time spent at specific location categories compared to a baseline period (January 3 - February 6, 2020), derived from anonymized location data.

^#^The comparisons are reported as mean differences.
